# Rapid identification of genomic structural variations with nanopore sequencing enables blood-based cancer monitoring

**DOI:** 10.1101/19011932

**Authors:** Jose Espejo Valle-Inclan, Christina Stangl, Anouk C. de Jong, Lisanne F. van Dessel, Markus J. van Roosmalen, Jean C.A. Helmijr, Ivo Renkens, Sam de Blank, Chris J. de Witte, John W.M. Martens, Maurice P.H.M. Jansen, Martijn P. Lolkema, Wigard P. Kloosterman

## Abstract

Somatic genomic structural variations (SVs) are promising personalized biomarkers for sensitive and specific detection of circulating tumor DNA (ctDNA) in liquid biopsies. However, affordable and fast identification of such SV biomarkers is challenging, which hinders routine use in the clinic. Here, we developed a novel approach - termed SHARC - for rapid discovery of somatic SVs as personalized tumor biomarkers. SHARC combines low coverage cancer genome sketching by using Oxford Nanopore sequencing with random forest classification and a dedicated filtering pipeline to enrich for somatic SVs. Our method leverages the real-time and long-read capabilities of nanopore sequencing to identify somatic SV breakpoints at nucleotide resolution from a tumor tissue biopsy within three days. We applied SHARC to tumor samples of high-grade ovarian and prostate cancer and validated on average ten somatic SVs per sample with PCR mini-amplicons. An accompanying method for SV breakpoint detection from liquid biopsies was devised based on digital PCR, enabling detection of cancer in a quantitative manner. Using this method, we retrospectively monitored treatment response in patients with metastatic prostate cancer. Our work demonstrates that SHARC forms a universal framework for rapid development of personalized biomarker assays for blood-based monitoring of any cancer type.

## Introduction

The pursuit of true precision medicine in oncology prompts us to improve the detection of cancer recurrence as well as accurate and fast monitoring of response to treatment. The current diagnostic paradigm for monitoring of cancer relies on imaging (CT/MRI scans), which inherently lacks sensitivity for both the initial diagnosis and to detect changes over time^1,2^. A promising approach to improve tumor detection is the use of liquid biopsies, that can be used to detect circulating cell-free DNA (cfDNA) from body fluids, such as blood, in a minimally invasive manner^3,4^. In patients with cancer, apoptotic and necrotic tumor cells shed fragmented DNA into the bloodstream (circulating tumor DNA, ctDNA), which contributes to the total levels of cfDNA^5^. A positive linear correlation between levels of ctDNA and tumor burden has been observed for multiple cancer types^6,7^. Furthermore, previous studies successfully utilized ctDNA to capture intra-tumoral genomic heterogeneity from all tumor locations within one patient^8,9^. In multiple cases, ctDNA analysis identified cancer recurrence months before clinical symptoms presented^10–12^.

The most commonly used biomarkers to differentiate between cfDNA from normal cells and ctDNA from tumor cells are somatic single nucleotide variants (SNVs). However, detection of somatic SNVs in liquid biopsies requires high coverage sequencing to reach sufficient sensitivity^3^. Moreover, the detection of these mutations in plasma in a sensitive manner requires the design of mutation specific detection assays or gene enrichment panels for ultra-deep sequencing of target loci^3,4^. Somatic genomic structural variation (SV) may serve as another type of tumor-specific biomarker to detect and quantify ctDNA with high sensitivity in liquid biopsies^11–14^. Most common solid cancers contain dozens to hundreds of somatic SVs^15,16^. Besides some recurrent gene fusions, like TMPRSS2-ERG in prostate cancer, the vast majority of these somatic SVs are patient- and tumor-specific^17^. Furthermore, SVs form a unique breakpoint junction between two joined DNA strands and can be validated by straightforward junction-spanning (quantitative) PCR assays^12^. Personalized detection of somatic SVs could be a powerful addition to point-mutation based assays for detection of ctDNA in patients with cancer.

Somatic SVs are commonly detected with short-read paired-end next generation sequencing (NGS). However, as SVs can be very large, short reads are less suited for SV detection, particularly in genomic regions with low complexity or in multi-breakpoint events^18–20^. Recently, long-read sequencing techniques from Oxford Nanopore Technologies (ONT) and Pacific Biosciences (PacBio) have emerged and their increased power for germline and somatic SV detection has been extensively demonstrated^19–23^. Moreover, ONT enables a short turnaround time and real-time data analysis for clinical applications^24^. We leveraged the long-read and fast sequencing capabilities of nanopore sequencing to develop SHARC (<u>S</u>tructural c<u>h</u>anges <u>a</u>s bioma<u>r</u>kers for <u>c</u>ancer). Our approach combines ONT-based low-pass whole genome sequencing (WGS) of tumor tissue with a computational method to detect somatic SVs without the need for sequencing a reference control sample. The computational part of SHARC combines random forest classification and germline SV filtering to enrich for somatic SVs, which subsequently enables the design of tumor-specific PCR-assays within three days. We demonstrate the clinical applicability of SHARC by tracking somatic SVs in longitudinal cfDNA samples of patients with metastatic prostate cancer.

## Results

### Detection of somatic structural variations from low coverage nanopore sequencing of tumor biopsies

To rapidly identify tumor-specific SV breakpoints, which can be used to track tumor dynamics in blood, we developed SHARC. The first step of the SHARC assay involves low coverage nanopore sequencing of genomic tumor-derived DNA (**Fig. 1A**). A single nanopore run on the MinION or GridION platforms typically generates between 5-15 Gbs of data^25^, corresponding to 1.5-5x coverage of the human genome. Next, the low coverage sequencing data are mapped to the reference genome followed by the detection of SV breakpoint junctions from split read mappings (**Fig. 1B**)^21^. Subsequently, a classification and filtering pipeline is applied to enrich for somatic SV breakpoints (**Fig. 1B**). Finally, PCR assays with mini-amplicons are designed to validate somatic SVs. SVs are confirmed as either somatic or germline by breakpoint PCR on tumor and corresponding lymphocyte DNA (**Fig. 1C**). Successful breakpoint PCR assays for somatic SVs detected with SHARC can then be utilized for ctDNA-based monitoring of treatment response and disease recurrence (**Fig. 1D**).

**Figure 1:**
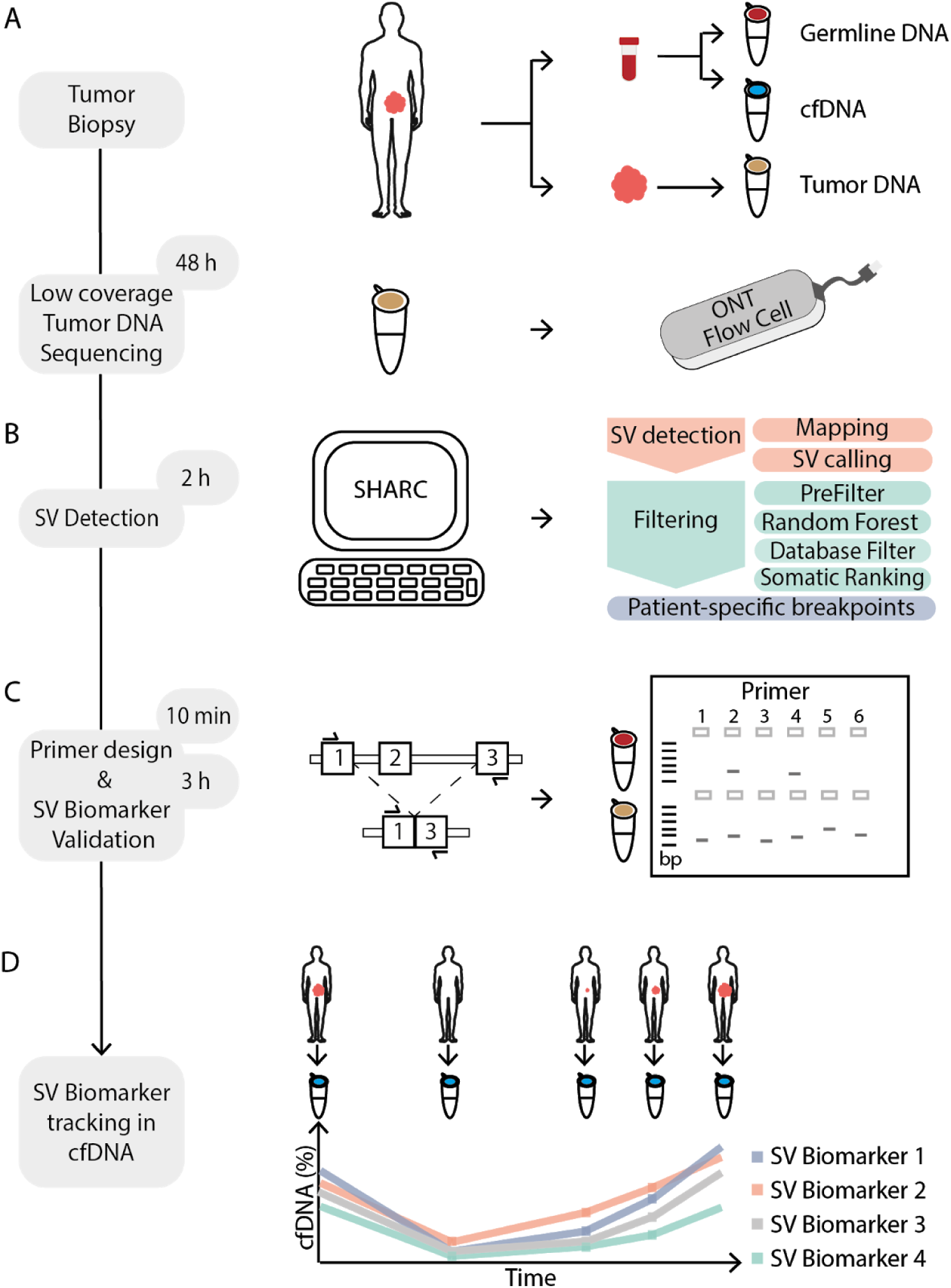
Schematic overview of SHARC. (**A**) (Needle) biopsy or resection from a tumor as well as blood are obtained from a patient at initial diagnosis. Germline DNA (red) and cfDNA (blue) isolated from blood and tumor DNA (brown) from tumor material. Tumor DNA is sequenced on one ONT flow cell. (**B**) Tumor-specific SV detection and filtering is performed with the bioinformatic SHARC pipeline. (**C**) SV-specific breakpoint spanning primers are designed. Breakpoint PCR with SV-specific primers is performed on germline and tumor DNA to confirm somatic SVs. (**D**) Somatic SVs are used as biomarkers and traced within cfDNA from a patient to monitor disease dynamics in a longitudinal manner.

### Establishment of a somatic SV reference set to test SHARC

To verify the ability of our pipeline to detect somatic SVs, we used genomic data from the melanoma cell line COLO829^26^ and the ovarian cancer organoid line HGS-3^27^. We utilized short-read WGS data from both lines (90x and 30x coverage for COLO829 and HGS-3, respectively) and matching reference samples (30x coverage in both cases) to establish two reference sets of somatic SVs (**Methods**). By using state-of-the-art SV detection pipelines^16,28^, we detected 92 and 295 somatic SVs in COLO829 and HGS-3, respectively. Additionally, we generated long-read nanopore sequencing data for COLO829 and HGS-3, reaching high coverages of 59x (COLO829) and 56x (HGS-3) (**Suppl. Fig. 1 and Suppl. Table 1**). To simulate low coverage long-read sequencing of tumor genomes, we randomly subsampled the nanopore sequencing reads to coverages of 4x, 3x and 2x. The subsampling was performed 20 times independently for each case, to mitigate the effect of chance on the subsampling and subsequent analysis.

Next, we tested the ability of SHARC to detect SVs from high and low coverage nanopore sequencing data. SHARC uses NanoSV^21^ to call SVs from the nanopore sequencing data. In total, 82,918 (COLO829) and 85,039 (HGS-3) SV candidates were called in the high coverage sets and an average of 12,886 (COLO829) and 11,284 (HGS-3) SV candidates were called in the low coverage subsets (**Suppl. Table 2)**. Based on the overlap with the somatic short-read reference set, raw SV calls were classified as somatic (true-positives) or non-somatic (false-positives). The vast majority of the raw SV calls in all the different coverage datasets were false-positives, on average 99.84% (range 99.81-99.9%, COLO829) and 99.55% (range 99.4-99.74%, HGS-3) **(Fig. 2A)**. Nevertheless, in the high coverage datasets, we identified 84 (91% of the short-read reference set, 0.1% of the total calls) and 219 (74% of the short-read reference set, 0.3% of the total calls) true-positive somatic SVs for COLO829 and HGS-3, respectively (**Fig. 2A and Suppl. Fig. 2A**). The fraction of true-positives with respect to the total number of SV calls was similar in the low coverage subsets (average of 0.18% and 0.5% for COLO829 and HGS-3, respectively) (**Fig. 2A**). Thus, we show that we can identify somatic SVs from both high and low coverage tumor-only nanopore sequencing data, despite the major fraction of SV calls being false-positives.

**Figure 2:**
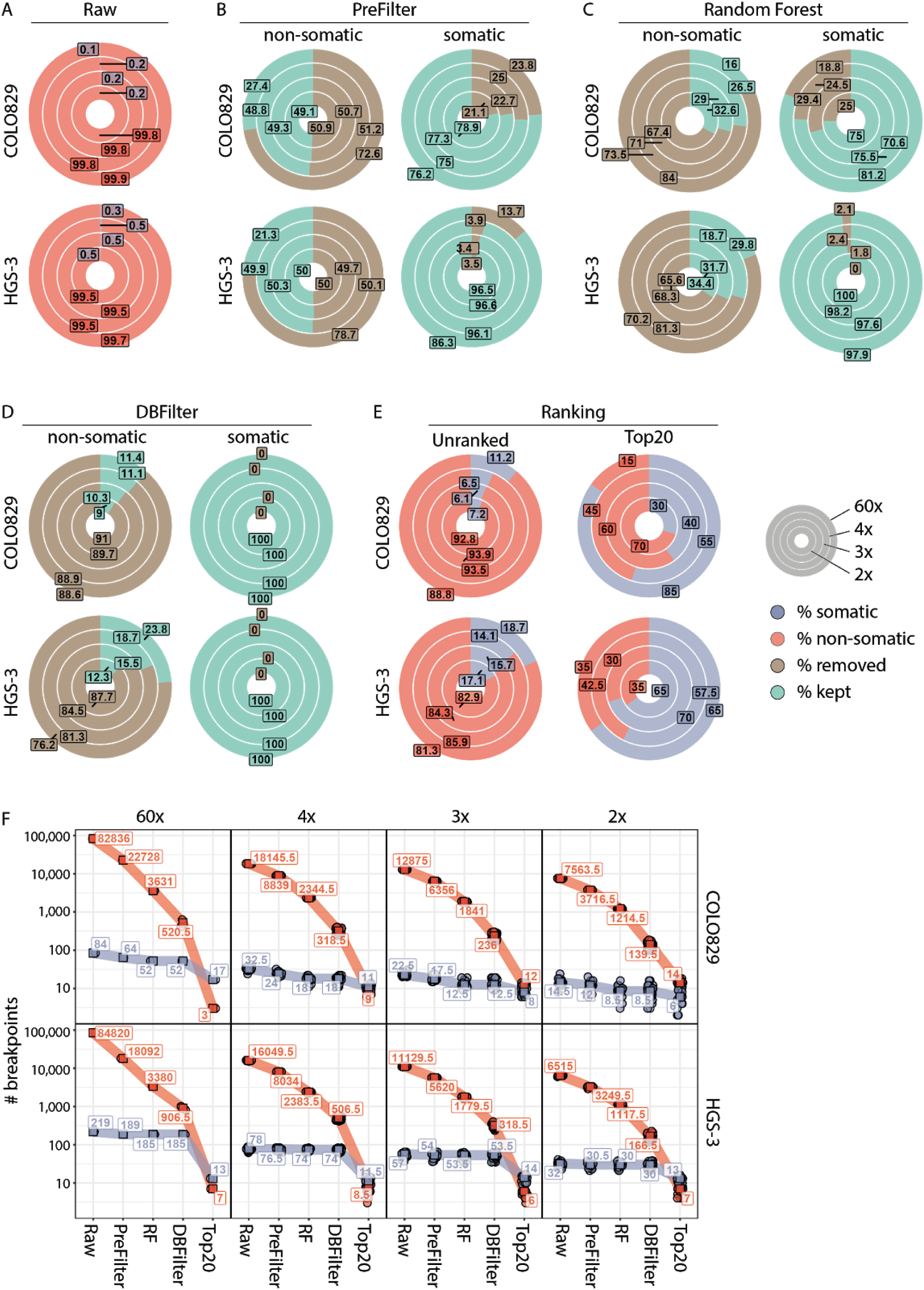
Detection of somatic SVs with the SHARC pipeline based on high and low coverage nanopore data. High coverage nanopore sequencing data from COLO829 (melanoma cell line) and HGS-3 (ovarian cancer organoid) were subsampled to low coverages. Outer circles represent the high coverage sets (59x for COLO829 and 56x for HGS-3) and inner circles represent low coverage subsets (4x 3x, 2x). **A**) Median percentage of non-somatic (red) and somatic (blue) breakpoints in the raw NanoSV calls for COLO829 (top) and HGS-3 (bottom). (**B**) Median percentage of non-somatic (left) and somatic (right) SV calls kept (green) or removed (brown) in the pre-filtering step for COLO829 and HGS-3. (**C**) Median percentage of non-somatic (left) and somatic (right) SV calls kept (green) or removed (brown) by the Random Forest SV classifier for COLO829 and HGS-3. (**D**) Median percentage of non-somatic (left) and somatic (right) SV calls kept (green) or removed (brown) by the database filtering for COLO829 and HGS-3. (**E**) Median percentage of non-somatic (red) and somatic SV (blue) calls in the complete SHARC output (left) and top 20 largest SVs (right) for COLO829 and HGS-3. (**F**) Total number of non-somatic (red) and somatic (blue) SV calls at each step of the pipeline for both COLO829 and HGS-3. In low coverage subsets, all data points are shown and the square box represents the median value. RF: Random forest; DBFilter: Database filter.

### Enrichment for true-positive SV calls from nanopore sequencing data

To reduce false-positive SV calls, we implemented a panel of filtering steps. First, we selected only “PASS” SV calls (based on default NanoSV filter flags^21^, **Methods**). Secondly, we excluded calls involving chromosome Y or the mitochondrial genome. Finally, we removed all insertions, since the exact inserted sequence cannot be accurately defined, thus hampering the final PCR assay development at a later step. As a result of these filtering steps, 72.6% (COLO829) and 76.2% (HGS-3) false-positive calls were removed in the high coverage sets (**Fig. 2B and Suppl. Table 2)**. For the low coverage sets, the filtering removed on average 50.9% (COLO829) and 49.9% (HGS-3) of false-positive calls (**Fig. 2B and Suppl. Table 2**). In contrast, the vast majority of somatic true-positive SV calls was maintained following SV filtering (on average 76.9% in COLO829 and 93.9% in HGS-3, **Fig. 2B**).

To further reduce the number of false-positive SV calls, we employed a random forest (RF) machine learning approach (**Methods**), similarly as previously described for SV calling of nanopore data^21^. We applied the RF classifier to the filtered high and low coverage subsets of COLO829 and HGS-3. For the high coverage sets, the RF labelled 84% (COLO829) and 81.3% (HGS-3) of false-positive SV calls as false (**Fig. 2C**). For the low coverage sets, on average 70.6% (COLO829) and 68% (HGS-3) of false-positive SV calls were labelled as false (**Fig. 2C**). In addition, in the high coverage sets, 81.25% (COLO829) and 97.88% (HGS-3) of true-positive SV calls were labelled as true. Similar percentages of true-positive SV calls were labelled as true in the low coverage sets, on average 73.7% (COLO829) and 98.6% (HGS-3) (**Fig. 2C**).

These results show that the RF classifier filters out the majority of false-positive breakpoints, while maintaining true-positive somatic SV calls. However, germline SV calls are also maintained at this step. Therefore, somatic true-positive SVs still represent a minor fraction of the total remaining SV calls: 0.3% COLO829 high coverage, 0.3% average COLO829 low coverage, 1% HGS-3 high coverage and 1% average HGS-3 low coverage **(Suppl. Fig. 2B)**.

To reduce the number of germline SVs, we implemented a blacklist filtering step. Therefore, the remaining SV calls were overlapped with two databases (DBFilter): (i) SharcDB, containing SV calls from nanopore sequencing of 12 samples (**Methods**) to remove both germline SVs and nanopore-specific false-positives, and (ii) RefDB, containing germline SV calls from 63 control samples previously sequenced using Illumina WGS in our group (**Methods**). Following this filtering step, 100% of true-positive somatic SV calls from both the COLO829 and HGS-3 high and low coverage sets were retained (**Fig. 2D**). In contrast, 88.6% (COLO829, high coverage), 76.2% (HGS-3, high coverage) and on average 89.9% (COLO829, low coverage) and 84.5% (HGS-3, low coverage) of remaining false-positive SV calls were filtered out (**Fig. 2D**). Due to this filtering, the fraction of true-positive somatic breakpoints in the remaining SV calls in the high coverage set increased to 11.2% (COLO829) and 18.7% (HGS-3) and to an average 6.6% (COLO829) and 15.6% (HGS-3) for the low coverage subsets (**Fig. 2E and Suppl. Fig. 2A**).

Finally, to further enrich for somatic SVs, we implemented a ranking method. Based on the observation that large SVs are more likely to be somatic than germline SVs (**Suppl. Fig. 3**), we ranked the remaining events based on length. Selecting the Top20 largest SV calls increased the percentage of true-positive somatic SVs to 85% (COLO829) and 65% (HGS-3) in the high coverage sets, and to on average 43% (COLO829) and 64.1% (HGS-3) in the low coverage sets (**Fig. 2E**).

Altogether, our SV filtering pipeline strongly enriches for true-positive somatic breakpoints and filters out the majority of false-positives and germline SVs. We demonstrate a total enrichment of true-positive somatic SV calls from 0.1% in the raw calls to 85% in the final Top20 ranked calls (17/20, COLO829, high coverage), 0.26% to 65% (13/20, HGS-3, high coverage), on average 0.18% to 41.7% (8.3/20, COLO829, low coverage sets) and on average 0.49% to 64.2% (12.8/20, HGS-3, low coverage sets) (**Fig. 2F and Suppl. Fig. 4**).

### Validation of SHARC in tumor tissue from patients with ovarian and prostate cancer

Next, we tested SHARC on four high-grade serous ovarian cancer (Ova1-4) and six prostate cancer (Pros1-6) samples. We sequenced tumor DNA on one nanopore flow cell per sample. The ovarian cancer samples and three prostate cancer samples (Pros1-3) were sequenced on commercial ONT flow cells. For the ovarian cancer samples, we started library preparation with minimally 1 µg of DNA. For the prostate cancer samples limited material was available, and we started library preparation with 250 ng of DNA. For one sample (Pros3), not enough sequencing data was produced to confidently detect somatic SVs and this sample was therefore excluded from all subsequent analyses (**Suppl. Table 1)**. Three additional prostate cancer samples (Pros4-6) were sequenced on ONT research prototype flow cells with higher sequencing sensitivity, thus requiring less DNA input material. In these cases, library preparation was started with an average of 108 ng (80-128 ng) of DNA and an average of 10 ng of library was loaded for sequencing (**Suppl. Table 1)**. We obtained an average sequence coverage of 2.3x (range: 1.8 - 4.0) (**Fig. 3A and Suppl. Table 1)** and average read lengths of 7.8 Kbp (range: 4.2-16.3 Kbp) (**Fig. 3B and Suppl. Table 1**). The sequencing throughput was not affected by the lower DNA input when using the high-sensitivity prototype flow cells. (**Suppl. Table 1)**.

**Figure 3:**
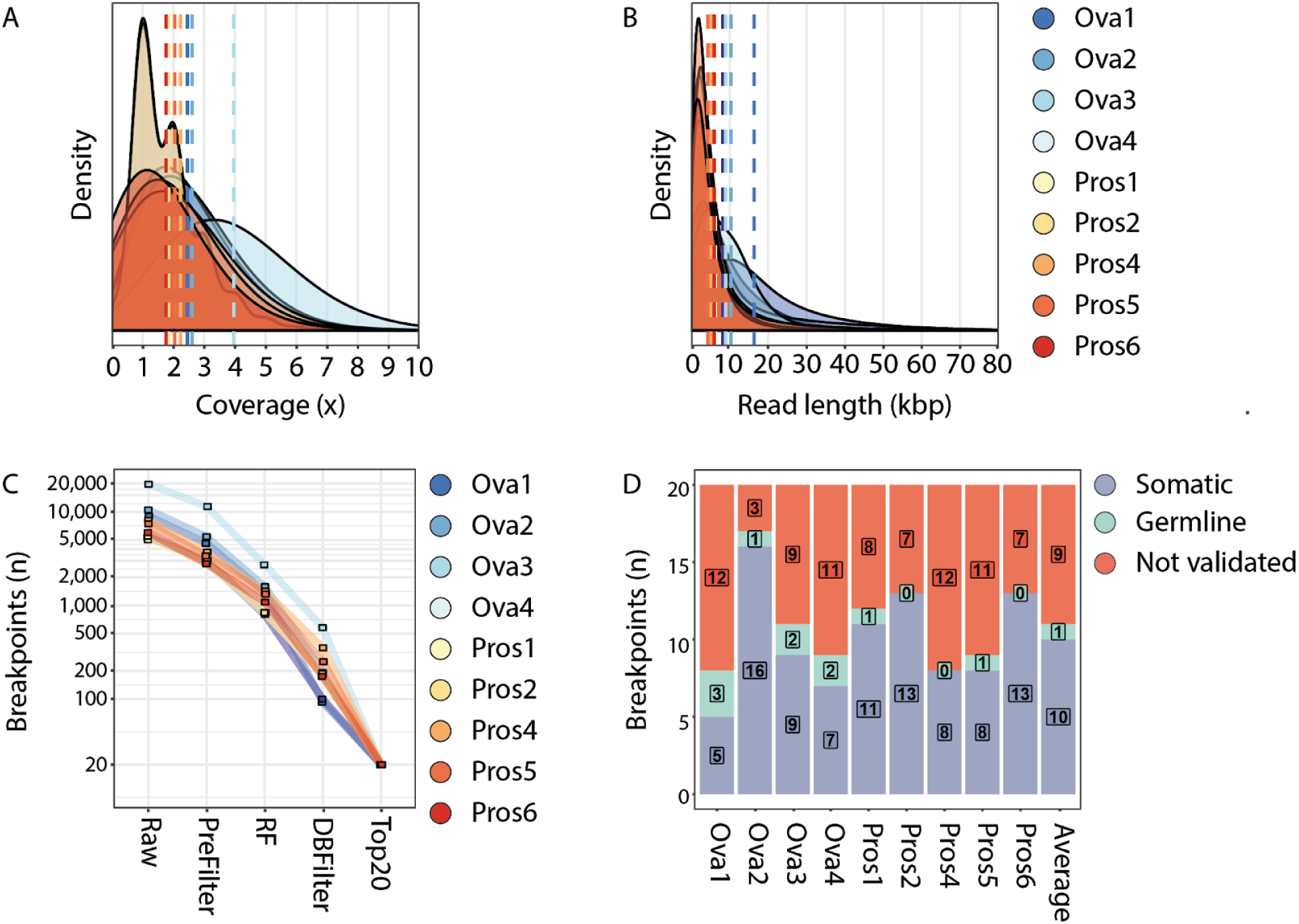
SHARC identifies and validates tumor-specific SV biomarkers from low-pass nanopore tumor sequencing data. Plots showing the distribution of (**A**) coverage and (**B**) read length for the nine tumor samples sequenced on one flow cell each. Dashed lines represent averages for each sample. (**C**) Total number of somatic SVs present at each of the steps throughout the SV calling and filtering pipeline. RF: Random forest; DBFilter: Database filter (**D**) The Top20 ranked breakpoints for each sample were tested by breakpoint PCR using tumor and germline DNA. Graph depicts the number of breakpoints validated as somatic (blue), germline (green) or breakpoints that could not be validated (red).

On average we identified 8,488 (range 5,024-19,695) raw SV calls in these samples (**Fig. 3C)**. Following the pre-filtering, the RF classification and the database filtering steps, an average 2.8% (range of 1.0%-4.4%) of SVs per sample were retained (**Fig. 3C**). We performed breakpoint PCR assays on lymphocyte and tumor DNA for the Top20 ranked SVs and validated an average of 50% (range 25-80%) somatic SVs per sample (**Fig. 3D**). We investigated the recall of validated somatic SVs at different timepoints during the sequencing run. We found that, on average, 81.6% (range 50-100%) of validated somatic SVs were already detected within the first 24 hours of sequencing (**Suppl. Fig. 5)**. This offers the opportunity to reduce the sequencing time, accelerating our SHARC assay with one day.

### Detection of SVs, identified by SHARC, in cfDNA from patients with ovarian and prostate cancer

To show the applicability of SHARC to detect clinically relevant biomarkers, we next tested if we could detect the validated somatic SVs in cfDNA of patients. Ascites fluid, which is known to contain cfDNA and ctDNA^29^ was available for Ova2 at time of recurrence. We extracted cfDNA from the ascites and tested all 16 validated somatic SVs by PCR. 100% of somatic SVs could be detected within the cfDNA from ascites (**Suppl. Fig**.**6**), and not in the germline or water controls. Next, we tested whether SVs identified and validated by SHARC, could be detected in cfDNA from blood. Therefore, we selected two patient-specific SVs for each of the four prostate cancer patients (Pros1, 4, 5 and 6) based on a high signal to noise ratio observed in qPCR assays for SV breakpoints (**Fig. 4A and Methods**).

**Figure 4:**
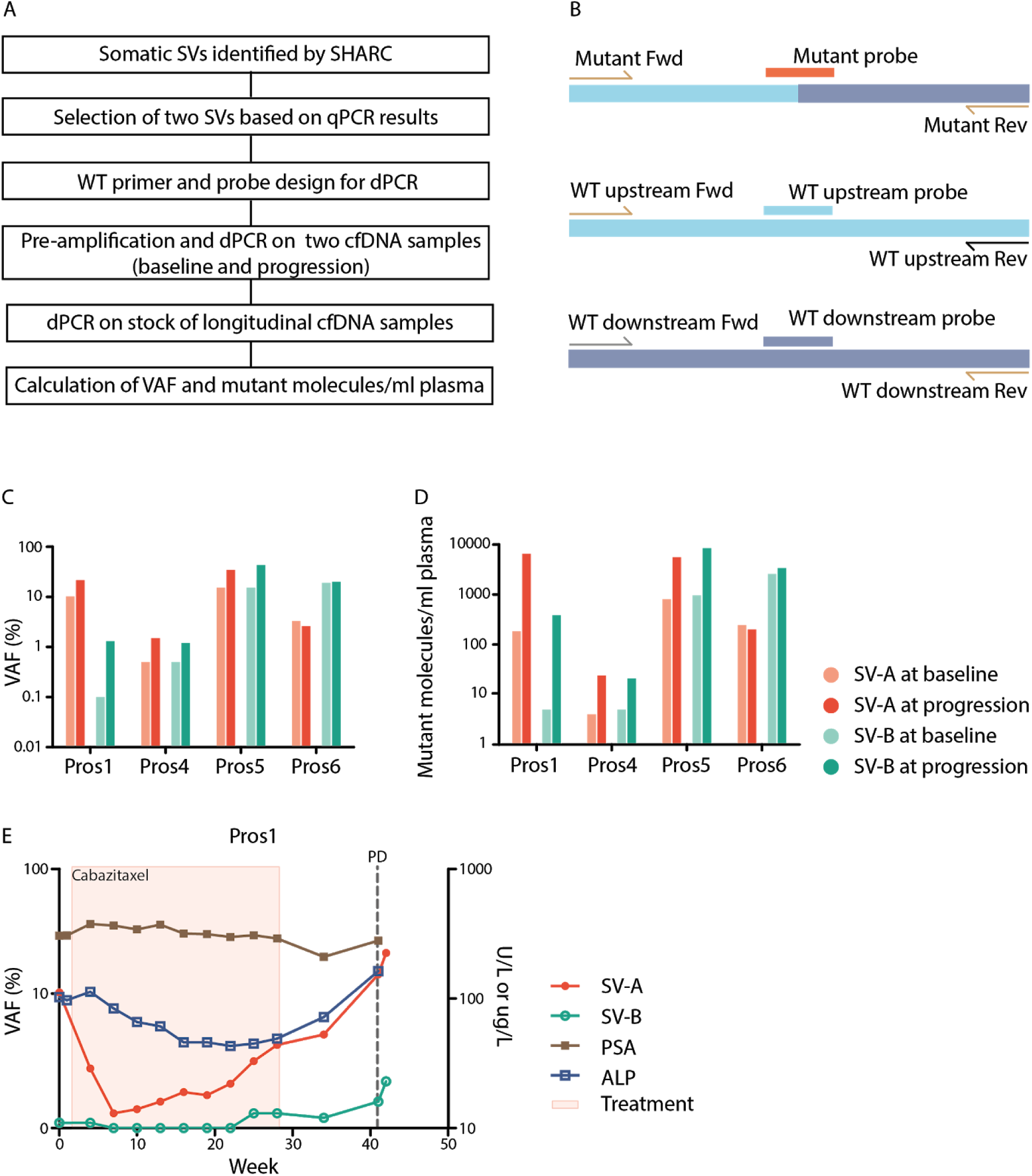
dPCR-based quantification of SVs in blood. (**A**) Schematic overview of quantification of tumor-specific SVs, identified by SHARC, in cfDNA from blood by using qPCR and dPCR. (**B**) Primer and probe design for dPCR. The wild-type upstream and wild-type downstream allele share each one primer with the mutant allele. Three probes with different fluorescents were designed to specifically detect the mutant allele or one of the wild-type alleles. (**C**) Detection of two tumor-specific SVs in cfDNA from blood from four patients with prostate cancer at baseline and at progression of disease with dPCR. Shown are VAF and (**D**) mutant molecules per mL plasma. (**E**) Quantification of SVs in longitudinal cfDNA samples from blood of patient Pros1. Graph depicts VAFs of SVs, treatment, laboratory parameters (prostate specific membrane antigen (PSA), alkaline phosphatase (ALP)) and clinical progression of disease (PD).

To enable sensitive and quantitative detection, we designed digital PCR (dPCR) assays for the eight selected SVs (**Fig. 4B**). For each SV, we aimed to design a probe for both wild-type alleles (up- and downstream) and for the mutant allele (across the breakpoint junction). For five SVs we could design an assay that quantified both the wild-type upstream and the wild-type downstream allele. For the three other SVs, primers/probes for only one of the wild-type alleles were designed, as appropriate primer design for the other allele was hindered by repetitive sequences at the target site. As the amount of cfDNA within one liquid biopsy is limited, we used a conditional breakpoint detection approach: (i) if dPCR on pre-amplified cfDNA (input pre-amplification: 0.2-1 ng cfDNA) confirmed the presence of the SV within cfDNA, (ii) then subsequent dPCR on non-preamplified cfDNA (stock cfDNA) (input dPCR: 5 ng cfDNA) was performed. The latter enabled calculation of both the variant allele frequency (VAF) and the number of mutant molecules per milliliter plasma (MM/mL plasma). First, we selected two timepoints per patient, one at baseline and one at progression of disease and confirmed the presence of all eight SVs with dPCR on pre-amplified cfDNA (**Suppl Fig. 7**). Thereafter, dPCR on the stock cfDNA successfully detected all SVs in the four patients, both in baseline and progression samples (**Fig. 4C and 4D**). Despite the fact that the VAF in pre-amplified cfDNA correlates to the VAF in stock cfDNA (r_s_ = 0.928), they should be considered two separate outcome measurements (regression coefficient = 0.72 ≠ 1) (**Suppl. Fig. 8A**). Moreover, VAF based on the wild-type upstream allele was highly similar to VAF based on the wild-type downstream allele in stock cfDNA (r_s_ = 0.996, regression coefficient = 1.05) (**Suppl. Fig. 8B**), suggesting no significant imbalances between the two sides of the breakpoint.

### Monitoring treatment response in patients with prostate cancer

In addition to the detection of SVs in cfDNA at baseline and progression of disease, we explored the capacity to use SVs, identified with SHARC, to monitor treatment response over time. To enable reliable response monitoring, measurements should be accurate and repeatable. As VAFs are ratios and in principle not influenced by technical variations between timepoints, we chose to report VAFs only. To verify the accuracy of dPCR, we performed two technical replicates for all pre-amplified samples of Pros5 and Pros6 and confirmed a high correlation of VAFs between the replicates (r_s_ = 0.987, regression coefficient = 0.918) (**Suppl. Fig. 8C**). Finally, we quantified the eight SVs of the four prostate cancer patients in the longitudinally collected samples from before, during and after treatment. For Pros1, SV-A shows the potential to improve response evaluation as its dynamics corresponds to the response to treatment with cabazitaxel and increases towards the end of treatment, resulting in the highest levels at clinical progression of disease (**Figure 4E**). These changes also seem to correlate with other blood biomarkers, including PSA and ALP. In addition, SV-B in Pros1 similarly correlates with response to treatment (**Figure 4E**). Also for Pros5 both SV-A and SV-B show clear changes over time correlating with clinical parameters, and Pros4 and Pros6 have less compelling dynamics of the detected SVs (**Suppl. Fig. 9A-C**).

## Discussion

Recent studies have described the use of somatic SVs for tracking tumor burden from liquid biopsies^11–14^. Although these studies showed the potential of this methodology, they lacked sufficient turn-around time to have personalized biomarkers ready before the initiation of patient treatment. This is a result of lengthy short-read WGS approaches for SV detection and an associated substantial number of false-positive somatic SVs, requiring laborious testing to validate SVs. To overcome these limitations, we developed SHARC. SHARC utilizes the real-time and long-read capabilities of nanopore sequencing combined with a machine learning approach to efficiently identify somatic SVs from tumor tissue within three days. The rapid nature and simple workflow of SHARC offers great potential for routine monitoring of cancer treatment response or detection of recurring disease. We demonstrate the applicability of SHARC to measure tumor burden by using a series of longitudinally gathered blood samples from metastatic prostate cancer patients. SHARC provides a universal method that can be applied for any cancer that is rich in somatic SVs.

Obtaining enough tumor material for DNA isolation is often a limiting factor for next-generation sequencing assays^12^. We show that nanopore sequencing and somatic SV detection is possible from limited amounts of DNA that can be extracted from a tumor needle biopsy. DNA input can be decreased even further to as little as 80 ng when using flow cells with increased sensitivity for DNA (research prototype flow cells provided by ONT). Thus, our assay can also be applied to patients with metastatic cancer that undergo a needle biopsy for diagnostic purposes, which is an important requisite for clinical viability.

Long-read sequencing is an excellent method for the detection of SVs at nucleotide resolution, even at low sequencing depth, because each long-read that bridges a breakpoint-junction provides direct information on the breakpoint position and sequence^21^. Sequencing of a tumor sample on a single GridION/MinION nanopore flow cell generates insufficient sequencing data to establish an accurate genomic profile. However, using the SHARC pipeline developed here, we efficiently enriched for somatic SVs despite very low coverage and without germline sequencing data. These assets make SHARC a cost-efficient assay for detection of somatic SVs as personalized cancer biomarkers. Furthermore, on average 50% of the SVs resulting from our pipeline are somatic, which minimizes the hands-on effort needed for validation purposes. For all analyzed tumors in this study, we identified at least five somatic SV biomarkers per patient, which is within the range described in previous work^11,13,30^. With expected increases in sequencing throughput from ONT sequencing, the performance from the SHARC pipeline will improve significantly. Furthermore, the use of cheap disposable flow cells (Flongle) could reduced assay costs to ⅕ of the current sequencing price of 800€^31^. The minimal costs of this assay would enable the broader application of such individualized SV tracing in patients with cancer.

We retrospectively applied SHARC to four patients with prostate cancer tracing levels of ctDNA by using SVs and compared tumor-dynamics to clinical biomarkers such as PSA and ALP. The quantitative measurement of SVs in ctDNA suggest that VAFs of SVs correlate with tumor load (Pros1 and Pros5). Moreover, the SVs would have indicated progression of disease earlier than PSA did in some patients (Pros1 and Pros 4). This clearly illustrates the potential clinical utility of quantifying ctDNA with SVs identified by SHARC to monitor response to treatment. However, further studies are needed to determine the universal applicability of the methods and the characteristics of the SVs that are particularly suitable for such strategies. Furthermore, SV-selection requires further improvement, to not only select tumor-specific SVs, but also SVs that represent the dominant disease clone. The approach of combining machine learning algorithms and experimental data could prove useful in optimizing this part of the assay. Larger prospective studies should confirm that indeed measuring SVs improves clinical decision making in patients with metastatic prostate cancer.

These data open the way to contemplate true individualized diagnostic platforms that are affordable in the current day clinical practice. The use of ONT or other desktop sequencing platforms combined with DNA printing machines that could provide individualized primers as needed, would enable such individualized approaches. The main barrier in curing cancer remains a lack of dynamic knowledge acquisition. We are very well aware of the dynamic response of cancer to treatment but lack the tools to monitor these changes in real time and thus generally respond to alterations too late for true treatment success. Individualizing disease monitoring could increase sensitivity to such levels that more intelligent treatment approaches could be envisioned.

## Materials and methods

### Patients

Tumor samples of four patients with high-grade serous ovarian cancer (OC) and six patients with metastatic castration resistant prostate cancer (PC) were used in this study. Patients with OC participated in the HUB-OVI study, in which tumor tissue and blood were obtained for organoid culture (tumor) and whole genome sequencing (WGS) (tumor and blood). Clinical data was extracted from the patient file in collaboration with the Dutch Cancer Registration. Patients with PC participated in both the CPCT-02 study (NCT01855477) and the CIRCUS study (NTR5732), in which tumor tissue from a metastatic lesion for WGS and longitudinal cfDNA samples were obtained. Longitudinal ctDNA quantification was performed for four patients with PC. Informed consent was obtained within all studies. Clinical data for patients with PC were collected in an electronic case report form (ALEA Clinical). All studies were performed according to the guidelines of the European Network of Research Ethics Committees (EUREC) following european, national and local law.

### DNA Isolation and nanopore sequencing

COLO829 (ATCC® CRL-1974™) cell line was obtained from the American Type Culture Collection (ATCC) and grown according to standard procedures as recommended by ATCC. DNA was isolated using a phenol chloroform protocol^32^. For some nanopore sequencing runs, DNA was sheared using g-tubes (Covaris). DNA was size selected on the PippinHT (Sage Science). Library preparation was performed using the Lib SQK-LSK109 kit (Oxford Nanopore Technologies) and DNA was then sequenced in 49 separate runs using R9.4 flow cells (Oxford Nanopore Technologies) on the MinION (44), GridION (3) and PromethION (2) instruments **(Suppl. Table 1)**.

HGS-3 organoid line was cultured following the ovarian cancer organoid culture protocol^27^. DNA was isolated by using a phenol chloroform protocol^32^. DNA was size selected on the PippinHT (Sage Science). Library preparation was performed using the Lib SQK-LSK109 kit (Oxford Nanopore Technologies) and DNA was then sequenced in 40 separate runs using R9.4 (23) and R9.5 (17) flow cells (Oxford Nanopore Technologies) on the MinION (35) and GridION (5) instruments **(Suppl. Table 1)**.

Tumor DNA from patients with ovarian cancer was isolated with the Genomic-tip kit (Qiagen), following the manufacturer’s protocol for tissue samples. DNA was prepared for nanopore sequencing with the Lib SQK-LSK109 (Oxford Nanopore Technologies). The library from one tumor sample was loaded on one revD (Ova1) or R9.4 (Ova2-4) flow cell (Oxford Nanopore Technologies). Sequencing was performed on a MinION (Ova2, Ova4) or GridION (Ova1, Ova3) instrument (Oxford Nanopore Technologies) **(Suppl. Table 1)**. Lymphocyte DNA for PCR validation assays was isolated from blood with the DNeasy Blood & Tissue Kit (Qiagen).

Tumor and germline DNA from patients with prostate cancer were obtained from a fresh frozen core needle biopsy of a metastatic lesion and blood, respectively. DNA was isolated on an automated setup with the QiaSymphony according to the supplier’s protocols (DSP DNA Midi kit for blood and DSP DNA Mini kit for tissue). In the context of the CPCT-02 study, WGS was performed by the Hartwig Medical Foundation, Amsterdam, The Netherlands^33^. Residual tumor DNA (80-250 ng) was used for nanopore sequencing. DNA was prepared for nanopore sequencing with the Lib SQK-LSK109 (Oxford Nanopore Technologies). The library from one tumor sample was loaded on one R9.4 (Pros1), revD (Pros2,3) or high-sensitivity research prototype (Pros4-6) flow cell (Oxford Nanopore Technologies). Sequencing was performed on a GridION instrument (Oxford Nanopore Technologies) (**Suppl. Table 1**).

### Illumina sequencing and analysis (COLO829 and HGS-3)

Short read WGS was obtained for matched tumor and normal DNA from the COLO829 cell line^34^ and the HGS-3 organoid line^27^.

SV calling was performed by using GRIDSS (v. 2.0.1)^28^ in joint calling mode (tumor+reference) for COLO829 and HGS-3 separately. Somatic SV calls were filtered as in^34^ (https://github.com/hartwigmedical/pipeline/blob/master/scripts/gridss_somatic_filter.R)

### SV calling and filtering pipeline

The SHARC pipeline is available through https://github.com/UMCUGenetics/SHARC. Mapping is performed in parallel for each FASTQ file by using minimap2 (v. 2.12)^35^ with settings “-x map-ont -a --MD”. The reference genome used is version GRCh37. Sorting and merging of BAM files was done by using sambamba (v. 0.6.5)^36^. SV calling was performed by using NanoSV (v. 1.1.2)^21^. Default NanoSV settings were used except a minimum read count of 2 (cluster_count=2) and minimum mapping quality of 20 (min_mapq=20).

VCFs are filtered by using the command ‘awk ‘$7 == “PASS” && $1 !∼ /(Y|MT)/ && $5 !∼/(Y|MT):/ && $5 != “<INS>“’’ to select PASS calls and remove insertions and SVs involving chromosomes Y or MT.

VCFs are then annotated with the distance to the closest single repeat element in the reference genome^37,38^, the closest gap element in the reference genome^38,39^, and the closest segmental duplication element in the reference genome^38^. These elements were taken from the UCSC genome browser (http://genome.ucsc.edu/)^38^, using the GRCh37/hg19 genome version.

We trained a random forest (RF) model to filter out false-positive SV calls from nanopore data, similarly as previously described^21^. We expanded the selection of input features for the RF, by including read length, SV calling features, and overlap with repeat features in the reference genome (**Suppl. Table 3**). We trained the classifier on the well-characterized NA12787 Genome in a Bottle (GIAB) sample^25,40,41^, for which high-quality germline SV call sets have been obtained by using Illumina^41^, PacBio^40^ and Nanopore^25^ sequencing. The GIAB SV truth set was generated by intersecting these three GIAB SV sets resulting in a set of 1,515 germline SVs. We used ⅔ of the GIAB truth set as a training set and ⅓ as a test set. We established a precision-recall curve from 100 bootstrapping runs (**Suppl. Fig. 3)**, where the training data were split into 90%-10% train-test subsets. Based on the precision-recall curve, we defined an operating point of 96% precision and 99.5% recall (**Suppl. Fig. 3)**. The final model was then re-trained on the whole training set and tested on the ⅓ test set. The performance on the test set was 95.1% precision and 99.6% recall, representing an accuracy of 97.2% (**Suppl. Fig. 3)**. SV candidates are classified as “true” or “false” based on this RF model.

We set up two databases of SV calls: (i) SHARCDB: containing raw NanoSV calls from nanopore sequencing data of 14 samples: COLO829-T, COLO829-BL, HGS-3, VCAP (prostate cancer cell line^42^), GIAB^25^, Ova1, Ova2, Ova3, Ova4, Pros1, Pros2, Pros4, Pros5 and Pros6 and (ii) RefDB: containing germline calls obtained from WGS short-read data of 59 controls: 19 blood controls from patients with ovarian cancer^27^, where germline SVs were called with Manta (v. 1.0.3)^43^ with default parameters and 40 healthy individuals (biological parents of individuals with congenital abnormalities)^44^ where germline SVs were called with Manta (v. 0.29.5)^43^ with default parameters.

SV calls from tumor samples are overlapped with those two databases by using VCF-explorer (https://github.com/UMCUGenetics/vcf-explorer).

Only samples classified as “true” by the RF model and that do not overlap with any sample in the databases qualify for primer design.

Primer design for filtered SV calls is automatized by using Primer3 (v. 1.1.4)^45^ with a product size range of 30-230 bp.

SVs with a successful primer design are ranked based on SV length and the 20 largest are selected for PCR validation.

### Breakpoint PCR

To validate SVs, breakpoint PCR with AmpliTaqGold (Applied Biosystems) was performed according to the manufacturer’s protocol. 10 ng primary tumor DNA (somatic) and 10 ng lymphocyte DNA (germline) per primer-pair were used as input. PCR products were loaded and visualized on a 2% agarose gel.

### cfDNA isolation

cfDNA was isolated from ascites fluid of Ova2 by using the QIAamp Circulating Nucleic Acid Kit (Qiagen) according to the manufacturer’s protocol. Plasma samples from patients with prostate cancer were obtained longitudinally during treatment in 3×10 ml CellSave preservative tubes (Menarini Silicon Biosystems, Huntingdon Valley, PA, USA) and processed within 96 hours as previously described^46^. Circulating DNA was isolated with the QIAsymphony® DSP Circulating DNA Kit (Qiagen) according to manufacturer’s protocol with some minor modifications^47^. All cfDNA samples were quantified by Qubit™ fluorometric quantitation (Invitrogen).

### Quantitative PCR

As primer specificity is essential for reliable interpretation of an end-point assay like dPCR, primers for the detection of structural variants were validated by quantitative PCR (qPCR) on whole genome amplified (WGA) tumor and germline DNA. In brief, qPCR was performed by using the CFX96 Touch™ Real-Time PCR Detection System (Bio-Rad Laboratories) and the final reaction mix consisted of 10 µL SensiFAST™ SYBR ® Lo-Rox mix (Bioline), 0.5 µM forward and reverse primers, 10 ng of WGA DNA and Ultrapure DNas/RNAse free H_2_O to bring up the reaction volume to 20 µL. The Cycle conditions were as follows: 14 cycles of 10s at 95°C and 30s at from 65-58°C (touchdown), followed by 20-40 cycles of 10s at 95°C and 30s at 60°C. In addition, a melt curve was generated from 56°C to 95°C to assess the generated PCR products. Based on qPCR results, two primer sets for the detection of SVs in each patient were selected for quantification by digital PCR (dPCR). Primer sets were excluded from use with dPCR when one of the following ocurred: >1 PCR product, Cq_germline_-Cq_tumor_ <5 and/or Cq_tumor_ > 20.

### DNA sonication and fragment size analysis

To mimic the length of cfDNA and improve DNA molecule partition, WGA DNA of both tumor and germline were sonicated to a peak size of ∼150 bp with the S220 Focused-ultrasonicator (Covaris) according to the manufacturer’s protocol. The sonication conditions were as follows; 200-250 ng WGA DNA (concentration determined by Qubit™ fluorometric quantitation) in 50 µL Ultrapure DNas/RNAse free H_2_O, Peak Incident Power: 175 W, Duty Factor: 10 %, Cycles per Burst: 200, Treatment Time: 280 s, Temperature: 7°C, and Water Level: 12. After sonication DNA fragment sizes were analyzed with the High Sensitivity DNA kit (Agilent Technologies) on the Bioanalyzer (Agilent Technologies) and the sample concentration was re-quantified by Qubit™ fluorometric quantitation (Invitrogen, Life Technologies, Carlsbad, CA, USA).

### Design of digital PCR assays for absolute quantification of SVs in cfDNA

To quantify SVs in cfDNA, dPCR was performed. First, the exact position of the breakpoint as determined by nanopore sequencing was validated. We used already available sequenced Illumina data from the CPCT-02 study (Pros1, Pros4, Pros5 and Pros6), but Sanger sequencing of the particular qPCR product could be used as well. To enable quantification of both mutant and wild-type alleles, additional primers for the detection of wild-type upstream (WT-U) allele and wild-type downstream (WT-D) allele of the breakpoint and fluorescent probes for both mutant and wild-type alleles were developed by using the Primer Express Software v3.0 (ThermoFisher) and the online tool Primer3Plus^45^. All primers and fluorescent probes (**Suppl. Table 4**) were ordered from Eurogentec.

### Pre-amplification of cfDNA

To enable sensitive detection of multiple SVs in limited amounts of cfDNA, two SVs per patient were pre-amplified with 0.2-1 ng of cfDNA. Pre-amplified tumor and germline DNA samples were used as respectively positive and negative control. Pre-amplification was performed by using 4 µL of TaqMan™ PreAmp Master Mix (cat.no: 4488593, Life Technologies), 2 µL primer pool (0.25 µM) consisting of SV forward (SV-F) and reverse (SV-R) primers and upstream (WT-U) and downstream (WT-D) wild-type primers, and 2 µL (cf)DNA for a total volume of 8 µL. Pre-amplification cycle conditions were: 10 min at 95°C followed by 14 cycles of 15 s at 95°C and 4 min at 60°C, and finally pause at 4°C. After the pre-amplification reaction, 72 µL of Ultrapure DNase/RNAse free H_2_O was added to each sample. Next, pre-amplified cfDNA was diluted 40x per 1 ng input, used for the pre-amplification, to prevent overloading of the dPCR chips.

### Absolute quantification of SVs in cfDNA with digital PCR

For the quantification of SVs in (cf)DNA, dPCR was performed with the Naica Crystal PCR system (Stilla Technologies) by using the following optimized reaction mix: 1 µL of diluted pre-amplified (cf)DNA sample, 5.6 µL PerfeCTa Multiplex qPCR ToughMix (Cat.No: 733-2322PQ, Quantabio). 0.25 µM probes (SV^FAM^, WT-U^HEX^, WT-D^CY5^), 0.75 µM of the SV forward (SV-F) and reverse primer (SV-R), 0.25 µM of the WT-U and WT-D primers, 0.1 µM Fluorescein (Cat.No: 0681-100G, VWR) and Ultrapure DNAse/RNAse free H_2_O to bring up the total volume to 28 µL. Samples were loaded onto Stilla Sapphire chips (Cat.no. C13000, Stilla Technologies) and dPCR was performed with the same cycle conditions as for the primer validation with qPCR. Median number of analyzable droplets was 21,357, inter quartile range 19,837-22,736. dPCR reactions were optimized with 10 ng sonicated tumor and germline WGA DNA. When an SV could be detected in pre-amplified cfDNA samples, a dPCR of all longitudinal cfDNA samples was performed on 5 ng of stock (no pre-amplification) cfDNA to enable absolute quantification of mutant molecules in plasma.

### Statistics

qPCR experiments were analyzed with Bio-Rad CFX Manager version 3.1. dPCR experiments were analyzed with Crystal Miner™ software, version 2.1.6 (Stilla Technologies). Thresholds for positive fluorescence were determined per primer pair based on positive and negative controls. Variant allele frequency (VAF) was calculated according to the following formula:

number of mutant molecules per µl in chip (as defined by Crystal Miner™ software) / (number of mutant molecules per µl in chip + number of wild-type molecules per µl in chip) * 100%.

Absolute number of mutant molecules per mL plasma was calculated as follows: number of mutant molecules per µl in chip * 28 µl (input in chip) / (used eluate/total volume of eluate * volume of plasma used for isolation).

To correct for zero values on a log scale, +1 was counted to every value and axes were corrected with -1. Spearman’s correlation coefficient was calculated for comparisons of VAF based on upstream wild-type allele vs downstream wild-type allele, two replicates and pre-amplified vs non-pre-amplified cfDNA samples. Corresponding slope was calculated by using linear regression analysis.

## Data Availability

Nanopore sequencing data is available in ENA and EGA as follows:
COLO829 cell line: ENA accession ERX2765498.
HGS-3 organoid line: EGA dataset accession EGAD00001005476
Patient material: EGA study accession EGAS00001003963

## Code availability

SHARC is available through https://github.com/UMCUGenetics/SHARC

## Data availability

Nanopore sequencing data is available in ENA and EGA as follows:

- COLO829 cell line: ENA accession ERX2765498.
- HGS-3 organoid line: EGA dataset accession EGAD00001005476
- Patient material: EGA study accession EGAS00001003963

## Author contributions

WPK and JEV-I conceived the study. JEV-I, SdB and MJvR performed bioinformatic experiments and JEV-I, SdB, MJvR, CS and WPK analyzed the data. IR performed nanopore sequencing. CS, IR, WPK and SdB performed wet lab experiments. CJdW, LFvD, MPL and ACdJ provided patient samples and clinical information. JCAH and AdJ performed cfDNA quantifications. JEV-I, CS, ACdJ, JCAH, LFvD, JWMM, MPHMJ, MPL and WPK interpreted data. JEV-I, CS and ACdJ wrote the manuscript. MPL and WPK edited the manuscript which was then reviewed and approved by all authors.

## Acknowledgements

The authors thank the former Kloosterman group at the UMC Utrecht and the Medical Oncology Department in the Erasmus MC for critical input. We thank Job van Riet for help with the design of dPCR assays. We thank Oxford Nanopore Technologies for providing the research prototype high sensitivity flow cells and the Utrecht Sequencing Facility for the nanopore sequencing. We thank all patients for providing the clinical specimens to perform this study. This work has been supported by KWF grants UU 2012-5710 and by funding from the Utrecht University to implement a single-molecule sequencing facility.

## References

1. Turkbey, B., Pinto, P. A. & Choyke, P. L. Imaging techniques for prostate cancer: implications for focal therapy. Nature Reviews Urology 6, 191–203 (2009).

2. Gerwing, M. et al. The beginning of the end for conventional RECIST — novel therapies require novel imaging approaches. Nature Reviews Clinical Oncology 16, 442–458 (2019).

3. Wan, J. C. M. et al. Liquid biopsies come of age: towards implementation of circulating tumour DNA. Nat. Rev. Cancer 17, 223–238 (2017).

4. Heitzer, E., Haque, I. S., Roberts, C. E. S. & Speicher, M. R. Current and future perspectives of liquid biopsies in genomics-driven oncology. Nat. Rev. Genet. 20, 71–88 (2019).

5. Schwarzenbach, H., Hoon, D. S. B. & Pantel, K. Cell-free nucleic acids as biomarkers in cancer patients. Nat. Rev. Cancer 11, 426–437 (2011).

6. Newman, A. M. et al. An ultrasensitive method for quantitating circulating tumor DNA with broad patient coverage. Nat. Med. 20, 548–554 (2014).

7. Abbosh, C. et al. Phylogenetic ctDNA analysis depicts early-stage lung cancer evolution. Nature 545, 446–451 (2017).

8. De Mattos-Arruda, L. et al. Capturing intra-tumor genetic heterogeneity by de novo mutation profiling of circulating cell-free tumor DNA: a proof-of-principle. Ann. Oncol. 25, 1729–1735 (2014).

9. Murtaza, M. et al. Multifocal clonal evolution characterized using circulating tumour DNA in a case of metastatic breast cancer. Nat. Commun. 6, 8760 (2015).

10. Diehl, F. et al. Circulating mutant DNA to assess tumor dynamics. Nat. Med. 14, 985–990 (2008).

11. Olsson, E. et al. Serial monitoring of circulating tumor DNA in patients with primary breast cancer for detection of occult metastatic disease. EMBO Mol. Med. 7, 1034–1047 (2015).

12. McBride, D. J. et al. Use of cancer-specific genomic rearrangements to quantify disease burden in plasma from patients with solid tumors. Genes Chromosomes Cancer 49, 1062–1069 (2010).

13. Leary, R. J. et al. Development of Personalized Tumor Biomarkers Using Massively Parallel Sequencing. Science Translational Medicine 2, (2010).

14. Klega, K. et al. Detection of Somatic Structural Variants Enables Quantification and Characterization of Circulating Tumor DNA in Children With Solid Tumors. JCO Precis Oncol 2018, (2018).

15. Li, Y. et al. Patterns of structural variation in human cancer. doi: 10.1101/181339

16. Priestley, P. et al. Pan-cancer whole-genome analyses of metastatic solid tumours. Nature (2019). doi: 10.1038/s41586-019-1689-y

17. Macintyre, G., Ylstra, B. & Brenton, J. D. Sequencing Structural Variants in Cancer for Precision Therapeutics. Trends Genet. 32, 530–542 (2016).

18. Dixon, J. R. et al. Integrative detection and analysis of structural variation in cancer genomes. Nat. Genet. 50, 1388–1398 (2018).

19. Chaisson, M. J. P. et al. Multi-platform discovery of haplotype-resolved structural variation in human genomes. Nat. Commun. 10, 1784 (2019).

20. Huddleston, J. et al. Discovery and genotyping of structural variation from long-read haploid genome sequence data. Genome Res. 27, 677–685 (2017).

21. Cretu Stancu, M. et al. Mapping and phasing of structural variation in patient genomes using nanopore sequencing. Nat. Commun. 8, 1326 (2017).

22. Nattestad, M. et al. Complex rearrangements and oncogene amplifications revealed by long-read DNA and RNA sequencing of a breast cancer cell line. Genome Res. 28, 1126–1135 (2018).

23. De Coster, W. et al. Structural variants identified by Oxford Nanopore PromethION sequencing of the human genome. Genome Res. 29, 1178–1187 (2019).

24. Jain, M., Olsen, H. E., Paten, B. & Akeson, M. The Oxford Nanopore MinION: delivery of nanopore sequencing to the genomics community. Genome Biol. 17, 239 (2016).

25. Jain, M. et al. Nanopore sequencing and assembly of a human genome with ultra-long reads. Nat. Biotechnol. 36, 338–345 (2018).

26. Pleasance, E. D. et al. A comprehensive catalogue of somatic mutations from a human cancer genome. Nature 463, 191–196 (2010).

27. Kopper, O. et al. An organoid platform for ovarian cancer captures intra- and interpatient heterogeneity. Nat. Med. 25, 838–849 (2019).

28. Cameron, D. L. et al. GRIDSS: sensitive and specific genomic rearrangement detection using positional de Bruijn graph assembly. Genome Res. 27, 2050–2060 (2017).

29. Husain, H. et al. Cell-Free DNA from Ascites and Pleural Effusions: Molecular Insights into Genomic Aberrations and Disease Biology. Mol. Cancer Ther. 16, 948–955 (2017).

30. Harris, F. R. et al. Quantification of Somatic Chromosomal Rearrangements in Circulating Cell-Free DNA from Ovarian Cancers. Sci. Rep. 6, 29831 (2016).

31. Gilpatrick, T. et al. Targeted Nanopore Sequencing with Cas9 for studies of methylation, structural variants and mutations. bioRxiv doi: 10.1101/604173

32. Quick, J. Ultra-long read sequencing protocol for RAD004 v3 protocols.io doi: 10.17504/protocols.io.mrxc57n

33. van Dessel, L. F. et al. The genomic landscape of metastatic castration-resistant prostate cancers reveals multiple distinct genotypes with potential clinical impact. bioRxiv doi: 10.1101/546051

34. Cameron, D. L. et al. GRIDSS, PURPLE, LINX: Unscrambling the tumor genome via integrated analysis of structural variation and copy number. bioRxiv doi: 10.1101/781013

35. Li, H. Minimap2: pairwise alignment for nucleotide sequences. Bioinformatics 34, 3094–3100 (2018).

36. Tarasov, A., Vilella, A. J., Cuppen, E., Nijman, I. J. & Prins, P. Sambamba: fast processing of NGS alignment formats. Bioinformatics 31, 2032–2034 (2015).

37. Benson, G. Tandem repeats finder: a program to analyze DNA sequences. Nucleic Acids Research 27, 573–580 (1999).

38. Haeussler, M. et al. The UCSC Genome Browser database: 2019 update. Nucleic Acids Res. 47, D853–D858 (2019).

39. Bailey, J. A. et al. Recent segmental duplications in the human genome. Science 297, 1003–1007 (2002).

40. Pendleton, M. et al. Assembly and diploid architecture of an individual human genome via single-molecule technologies. Nat. Methods 12, 780–786 (2015).

41. 1000 Genomes Project Consortium et al. A global reference for human genetic variation. Nature 526, 68–74 (2015).

42. Korenchuk, S. et al. VCaP, a cell-based model system of human prostate cancer. In Vivo 15, 163–168 (2001).

43. Chen, X. et al. Manta: rapid detection of structural variants and indels for germline and cancer sequencing applications. Bioinformatics 32, 1220–1222 (2016).

44. Middelkamp, S. et al. Prioritization of genes driving congenital phenotypes of patients with de novo genomic structural variants. bioRxiv doi: 10.1101/707430

45. Untergasser, A. et al. Primer3--new capabilities and interfaces. Nucleic Acids Res. 40, e115 (2012).

46. van Dessel, L. F. et al. Application of circulating tumor DNA in prospective clinical oncology trials - standardization of preanalytical conditions. Mol. Oncol. 11, 295–304 (2017).

47. van Dessel, L. F. et al. High-throughput isolation of circulating tumor DNA: a comparison of automated platforms. Mol. Oncol. 13, 392–402 (2019).

